# “Subtherapeutic concentrations of first-line antitubercular agents in pediatric patients and its association with tuberculosis treatment outcome: protocol for a systematic review and meta-analysis”

**DOI:** 10.1101/2020.05.19.20107177

**Authors:** Jorge Chachaima-Mar, Diana Sánchez-Velazco, Cesar Ugarte-Gil

**Author notes:** Correspondence Jorge Emerson Chachaima Mar Jr. Alfonso Ugarte 376, Lima, Perú 15102.

## Abstract

Pediatric tuberculosis is a neglected disease that is receiving more attention lately. Some studies found that serum levels of first line antituberculosis drugs do not reach reference concentrations in children. However, these reference ranges were validated in an adult sample. Thus, we do not know if subtherapeutic concentrations of antitubercular agents in children are associated with negative outcomes.

**Objective:** To estimate the association between subtherapeutic concentrations of first-line antitubercular drugs with clinical outcomes of treatment.

**Methods:** We propose to do a systematic review and meta-analysis. In order to do so, we will perform an electronic search in Medline, SCOPUS, Web of Science and Global Index Medicus. There will be no restriction of language nor date of publication. First, we will screen titles and abstracts; then we will screen through the full text of the article. Both phases will be done by 2 independent authors. Data extraction will be performed using a data abstraction form by two independent authors. The quality of the studies will be checked with standardized tools according to the design of the study, and will also be performed by duplicate. We will present the main characteristics of each included study through tables. The heterogeneity between studies will be assessed through the I^2^ statistic. If appropriate, we will use the random-effects model to calculate the pooled estimate. We will evaluate the publication bias through visual inspection of the funnel plot and Egger’s test. Pre-arranged subgroup and sensitivity analysis will be performed.

**Results:** We will publish the results of this systematic review in a peer-reviewed journal.

**Conclusions:** This systematic review will provide up-to-date evidence regarding serum concentration in pediatric patients and its association with outcomes. With the analysis we plan, we will offer important recommendations regarding the dosage of the first line antitubercular agents in children, and the modifications that may be needed.

**Conflicts of interest:** All the authors declare to have no conflict of interest.

**Funding:** This study did not receive funding from the public, commercial or not-for-profit sectors.

## BACKGROUND

With a quarter of the world’s population infected latently, tuberculosis still continues to be a public health challenge(1). Global eradication of this disease is the long term goal, that is why the World Health Organization(WHO) proposed the “END TB” strategy, with which WHO hopes, besides other objectives, to decrease by 95% deaths by 2030(2).

However, there is a long road ahead to accomplish these proposed goals. Pediatric tuberculosis is a clear example because it has been a neglected disease for a long time(3,4) until recent years that it has gained more importance(5). Currently, tuberculosis is the leading infectious cause of death in children(6), with approximately 200 000 deaths per year(7).

Generally speaking, this age group develops a paucibacillary disease which means that they often get negative sputum despite having an active disease(8,9); this makes the diagnosis more difficult and easy to overlook leading to the sub estimation of the real problem(10). In 2012, the WHO published estimates of pediatric tuberculosis for the first time with the number of new cases being 500000 per year(11). But, in 2015, the WHO report duplicated the estimate, being 1 000 000 new cases of pediatric tuberculosis every year(12). The reason behind this big change was the adjustment done taking into account the underreporting and under-registration of pediatric cases of tuberculosis(10).

Even though pediatric cases of tuberculosis have decreased worldwide, in high burden countries they can constitute 20% of the total number of cases(13). These countries usually have a population pyramid with a broad base, which increases the number of pediatric patients with tuberculosis(3). Furthermore, these countries often have a weak health system which makes it more difficult to attack this problem(7). In addition to these circumstances, children develop more often severe forms of the disease like meningitis or miliary tuberculosis(8,14,15); these could even constitute 80% of the cases in children under 3 years of age(14).

Even with treatment, mortality in children is high(16–18) with 20% in extrapulmonary tuberculosis(19). The risk of death is even higher in children with other comorbidities such as the Human Immunodeficiency Virus(HIV), (16,20–23), which can be present in up to 48% of children with tuberculosis (24,25).

Recommended doses by the WHO for the treatment of pediatric tuberculosis has been changing (26–28). There are two main reasons for this variation of the doses over time. Firstly, the actual treatment regimen against tuberculosis is based on 4 drugs(rifampicin, isoniazid, ethambutol, and pyrazinamide) which have demonstrated to be highly effective for the treatment of tuberculosis that are sensitive to such drugs(29–31). Most of the studies for this regime were done in adults, as clinical trials often exclude children for the difficulty to define a pediatric case of tuberculosis(5) limiting our knowledge of the pharmacokinetic properties of antituberculosis agents.

Another key point is that pharmacokinetic parameters in children of antitubercular antibiotics are not completely elucidated. These parameters have always been considered important to predict the efficacy of the tuberculosis treatment(32,33), being two of the most important peak concentration(Cmax) and the area under the curve(AUC) (33–37). The problem with these two parameters is the multiple sampling needed and the cost of each analysis. That is why a simplified sampling at the concentration of the measurement at the second hour(C2h) and at the sixth hour(C6h) was proposed. Nowadays, the guideline of the American Thoracic Society mentions that a sampling at the second hour and one more at the sixth hour could be done in patients with slow response to treatment or certain comorbidities like obesity(38).

What is well-known is that pediatric patients have a different pharmacodynamic profile which differs from the adult one (39), which may influence the serum concentration of the medications(40). Moreover, the reference range for the therapeutic concentration of antituberculosis drugs used for children and adults, comes from a study with an adult sample(41).

With these key points in mind, we can understand why the changes to the pediatric dosage of anti tuberculosis antibiotics. In the first edition of the guidelines of pediatric tuberculosis, the WHO extrapolated the dosage from the adults to the children population(42). The only exception was ethambutol because there was evidence that the Cmax in children did not reach the same levels as adults(28,43,44). The following years after the publication of the 2006 guideline, many studies evidence that pediatric patients did not reach recommended serum levels of antitubercular drugs(26,45–49). With the new evidence, the WHO decided to change the pediatric dosage in 2010(27,48), increasing the doses of rifampicin, isoniazid and pyrazinamide; these changes were later ratified in the second guideline of the year 2014(26).

However, even with the latest recommended doses there is contradictory evidence, some studies found inadequate serum drug concentrations, meanwhile, others found them to be adequate(50–53). To administer the right dose to pediatric patients with the maximum benefit and the least disadvantage, we need to know if the actual reference concentration range of the antituberculosis drugs is associated with the outcomes of the treatment. That is why we propose to do a systematic review of the existing evidence of the serum concentrations of the first-line antitubercular drugs in pediatric patients diagnosed with active tuberculosis and its effect on the clinical outcome.

## METHODS

### Study Design

This systematic review will be performed according to the Preferred Reporting Items for Systematic Review and Meta-Analysis(PRISMA)(54).

### Eligibility Criteria

#### Participants

- We will include pediatric patients (18 or younger) who are receiving first-line antitubercular agents for fully sensitive active tuberculosis. The diagnosis of tuberculosis could be done through microbiological tests (sputum smear, positive culture, molecular test), radiological images or clinical criteria. There will not be any dose restriction.

#### Exposition

- We will consider those patients with subtherapeutic concentrations of the first-line antituberculous drugs.

#### Comparison

- Patients with therapeutic drug concentration of the same drugs.

#### Types of Studies

- We will include experimental and observational studies including cross-sectional, case-control, cohorts, randomized and non-randomized clinical trials.

### □Exclusion Criteria

- We will exclude the following publication types: case reports, case series, letter to the editor, editorial, narrative review and systematic reviews.
- We will exclude patients with tuberculosis that are resistant to one or more antitubercular drugs.
- We will exclude those studies whose participants are only healthy children.
- We will exclude pharmacokinetic/pharmacodynamic modeling and simulation.
- We will exclude any study assessing latent tuberculosis.

#### Literature Search and Data collection

- The databases selected for this study are PubMed, Web Of Science, Global Index Medicus and SCOPUS. We will retrieve all articles to a Microsoft Excel 2016 sheet. The complete search strategy can be found in Appendix 1(Supplementary Materials).
- Then, one author will delete duplicates. Two authors will screen through titles and abstracts looking for relevant studies. Then, once more, two independent authors will look into the full text of previously selected articles. Any disagreement will be solved by discussing it or consulting a third author. We will record the reason for exclusion in the full-text phase and summarize all the information and the number of articles in every phase in a PRISMA flow diagram.
- We will also search the abstract books of the last 3 years of the “Union World Conference on Lung Health”. This manual search will be done by two independent authors.

#### Data extraction

- Using a previously developed data extraction form in Google Forms, then, a pilot testing will be conducted with some of the included articles. With the final version of the data extraction form, two independent authors will carry out data collection. The data will be exported to a Microsoft Excel sheet. Disagreements will be resolved by discussion until arriving at a consensus, and if that option fails, by consulting a third author.
- The data extracted will include:
  - Study details: First author, corresponsal author, article title, country, year of publication, funding source
  - Sample size(number of participants in each group/cohort/arm), inclusion criteria and exclusion criteria, diagnosis criteria or case definition, age range.
  - Study methodology: fasting time before administration of the drugs, hours when the serum sampling took place, how much time after starting the tuberculosis treatment did the sampling took place.
  - Treatment details: pill presentation(standard or fixed-dose combination), drugs doses, the reference range for the serum concentrations of the drugs or the pharmacokinetic parameters, number and proportion of patients with therapeutic serum concentration, including peak serum concentration (Cmax), concentration at 2 hours (C2h) and 6 hours (C6h), area under the curve (AUC), and other parameters such as: time to reach Cmax(tmax), peak serum concentration over minimum inhibitory concentration (Cmax/MIC), area under the curve over minimum inhibitory concentration (AUC/MIC).
  - Treatment outcomes: mean and standard deviation, or number and proportion of the participants with the concentration values within the reference range. We will assess treatment outcomes defined by the WHO[26] including treatment success(cure, treatment completed) treatment failed, death, lost to follow-up. We will evaluate acquired drug resistance, relapse and time to sputum culture or smear conversion as the additional outcomes.
  - Association between concentration and the treatment outcome

If we find any article where our population is mixed with any or if any required data is missing or unclear, we will try to contact the correspondent author. In case we do not have an answer from the corresponding author and the article does not report the results of our relevant population separately, we will exclude the study.

If data from a study was reported in more than one article, we will only select the article with the most complete data for our research purposes.

Serum concentration of the antitubercular drugs will be extracted in two ways, as a numerical variable and as a dichotomous variable. For the latter, we will divide the variable into subtherapeutic and therapeutic concentrations. We will also record the thresholds used to divide the two groups.

For all outcomes, we will also divide into two groups, ending with a dichotomous variable: successful treatment (including completed treatment or cured) and unsuccessful treatment(including treatment failed, died, relapse, lost to follow-up).

#### Risk of Bias and Methodological quality of Studies

We will assess the risk of bias by duplicate and independently using different tools according to the design of the study:

- Risk of Bias(ROB) from the Cochrane collaboration(55) for the randomized clinical trials.
- ROBINS-I tool for non-randomized clinical trials(56)
- The “National Heart Lung and Blood Institute” scales for cross-sectional, cohort and case-control studies(57).

If any discrepancy appears, it will be discussed until reaching a consensus, if that strategy fails, a third author will take the final decision. We will present a table with the results of this assessment.

### Statistical analysis

We will present data from included studies using tables summarizing the main data. We will also provide a narrative synthesis of the evidence found, and we will present tables with the most important data from the included studies. We will conduct all statistical analyses using Stata 15.1. We will assess heterogeneity with the I^2^ statistic. We will consider >75% or greater values as “high heterogeneity” (58,59). Assuming a high heterogeneity, we will pool the estimates through the random-effects model. We will generate a forest plot with the result of the meta-analysis. If feasible, we will assess publication bias through the Egger’s test and visually with the funnel plot.

If possible, subgroup analysis will be performed: age(<1 year old, <3 years old), dosage form(fixed vs standard), comorbidities(HIV, malnutrition, isoniazid acetylator status). Sensitivity analysis will be performed according to the classification of risk of bias.

## Data Availability

Not applicable, as this is a systematic review protocol.

## Notes

### Competing Interest Statement

The authors have declared no competing interest.

### Author Declarations

All information used will be from the public domain, thus, there is no need for a ethics committee approval

### Summary of Updates

We added a database(Global Index Medicus) for the electronic search. Everything else stays the same.

