## Supplementary Material for "“Subtherapeutic concentrations of first-line antitubercular agents in pediatric patients and its association with tuberculosis treatment outcome: protocol for a systematic review and meta-analysis”"

**Appendix 1**

**Search strategy**

| **DATABASE** | **SEARCH STRATEGY** | **FINAL** |
| --- | --- | --- |
| **Pubmed/Medline** | #1: ("Infant"[mh] OR "Adolescent"[mh] OR "Child"[mh] OR infant*[tiab] OR newborn*[tiab] OR neonat*[tiab] OR child*[tiab] OR kid[tiab] OR toddler*[tiab] OR adolescent*[tiab] OR teenager*[tiab] OR youth*[tiab] OR pubert*[tiab] OR paediatric*[tiab] OR pediatric*[tiab] OR school*[tiab] OR kindergar*[tiab] OR patient[tiab] OR patients[tiab])  #2 :(( (Rifampicin[tiab] OR rifampin[tiab] OR rifampin[mh]) OR ( isoniazid[mh] OR isoniazid[tiab]) OR (ethambutol[mh] OR ethambutol[tiab] OR pyrazinamide[mh] OR pyrazinamide[tiab]) OR antitubercul*[tiab] OR "Antibiotics, Antitubercular"[mh] OR antimycobacterial*[tiab]))  #3: (((concentration*[tiab] OR monitoring[tiab] OR level*[tiab]) AND (serum[tiab] OR plasma[tiab])) OR pharmacokinetic*[tiab] OR “therapeutic drug monitoring”[tiab] )  #4: ((case report[ti] OR case reports [pt] OR editorial[tiab] OR letter[ti] OR animal*[ti] OR Animals, Laboratory[Mesh] OR Animal Experimentation[Mesh] OR "Rodentia"[Mesh] OR rats[ti] OR rat[ti] OR mouse[ti] OR mice[ti] ))  Final: ((#1 AND #2)AND #3) NOT #4 | (("Infant"[mh] OR "Adolescent"[mh] OR "Child"[mh] OR infant*[tiab] OR newborn*[tiab] OR neonat*[tiab] OR child*[tiab] OR kid[tiab] OR toddler*[tiab] OR adolescent*[tiab] OR teenager*[tiab] OR Youth*[tiab] OR pubert*[tiab] OR paediatric*[tiab] OR pediatric*[tiab] OR school*[tiab] OR kindergar*[tiab] OR patient[tiab] OR patients[tiab]) AND (((Rifampicin[tiab] OR rifampin[tiab] OR rifampin[mh]) OR ( isoniazid[mh] OR isoniazid[tiab]) OR (ethambutol[mh] OR ethambutol[tiab] OR pyrazinamide[mh] OR pyrazinamide*[tiab]) OR antitubercul*[tiab] OR "Antibiotics, Antitubercular"[mh] OR antimycobacterial*[tiab])) AND (((concentration*[tiab] OR monitoring[tiab] OR level*[tiab]) AND (serum[tiab] OR plasma[tiab])) OR pharmacokinetic*[tiab] OR “therapeutic drug monitoring”[tiab] ) ) NOT ( ((case report[ti] OR case reports [pt] OR editorial[tiab] OR letter[ti] OR animal*[ti] OR Animals, Laboratory[Mesh] OR Animal Experimentation[Mesh] OR "Rodentia"[Mesh] OR rats[ti] OR rat[ti] OR mouse[ti] OR mice[ti] )) ) |
| **Scopus** | #1: TITLE-ABS-KEY(infan* OR newborn* OR baby OR neonate* OR child* OR kid OR toddler* OR adolescent* OR teenager* OR youth* OR puberty* OR p?ediatric* OR school* OR kindergar* OR patient OR patients)  #2: TITLE-ABS-KEY ((( ((rifampin* OR rifampicin*) OR ( isoniazid) OR (ethambutol OR pyrazinamide)) OR (antitubercul*) OR antimycobacterial*))  #3: TITLE-ABS-KEY(((concentration* OR level* OR monitoring) AND (serum OR plasma)) OR pharmacokinetic* OR “therapeutic drug monitoring”))  #4: TITLE( editorial OR case report* OR “animal experimentation*” OR animal* OR mouse OR mice OR rat ) OR KEY( editorial OR case report* OR “animal experimentation*” OR animal* OR mouse OR mice OR rat ) OR DBCOLL ( *medl)*  Final: ((#1 AND #2) AND #3) AND NOT #4 | ( ( ( TITLE-ABS-KEY ( infan* OR newborn* OR baby OR neonate* OR child* OR kid OR toddler* OR adolescent* OR teenager* OR youth* OR puberty* OR p?ediatric* OR school* OR kindergar* OR patient OR patients ) ) AND TITLE-ABS-KEY ( rifampin* OR rifampicin* OR isoniazid OR ethambutol OR pyrazinamide OR ( antitubercul* ) OR antimycobacterial* ) AND ( TITLE-ABS-KEY( ((concentration* OR level* OR monitoring) AND (serum OR plasma)) OR pharmacokinetic* OR “therapeutic drug monitoring”) ) AND NOT ( TITLE ( editorial OR case AND report* OR "animal experimentation*" OR animal* OR mouse OR mice OR rat ) OR KEY ( editorial OR case AND report* OR "animal experimentation*" OR animal* OR mouse OR mice OR rat ) OR DBCOLL ( medl ) ) ) ) |
| **Web Of Science** | #1 TS=( (infan* OR newborn* OR baby OR neonat* OR child* OR kid OR toddler* OR adoles* OR teenager* OR Youth* OR pubert* OR p$ediatric* OR school* OR kindergar* OR patient?))  #2: TS= (rifampin* OR rifampicin* OR isoniazid OR ethambutol OR pyrazinamide OR antitubercul* OR antimycobacterial* )  #3: TS=(((concentration* OR level* OR monitoring) AND (serum OR plasma)) OR pharmacokinetic* OR “therapeutic drug monitoring”)  #4: TI=( (case report* OR editorial OR letter OR animal* OR “animal experimentation*” OR rodent* OR rat* OR mouse OR mice)) OR AK=(case report* OR editorial OR letter OR animal* OR “animal experimentation*” OR rodent* OR rat* OR mouse OR mice) OR KP=(case report* OR editorial OR letter OR animal* OR “animal experimentation*” OR rodent* OR rat* OR mouse OR mice)  Final: ((#1 AND #2) AND #3) NOT (#4) | (((TS=( (infan* OR newborn* OR baby OR neonat* OR child* OR kid OR toddler* OR adoles* OR teenager* OR Youth* OR pubert* OR p$ediatric* OR school* OR kindergar* OR patient?)) ) AND (TS= (rifampin* OR rifampicin* OR isoniazid OR ethambutol OR pyrazinamide OR antitubercul* OR antimycobacterial* )) ) AND (TS=(((concentration* OR level* OR monitoring) AND (serum OR plasma) ) OR pharmacokinetic* OR “therapeutic drug monitoring”) ) ) NOT (TI=( (case report* OR editorial OR letter OR animal* OR “animal experimentation*” OR rodent* OR rat* OR mouse OR mice)) OR AK=(case report* OR editorial OR letter OR animal* OR “animal experimentation*” OR rodent* OR rat* OR mouse OR mice) OR KP=(case report* OR editorial OR letter OR animal* OR “animal experimentation*” OR rodent* OR rat* OR mouse OR mice)) |
| **Global Index Medicus** | 1. ((mh:Infant OR mh:Adolescent OR mh:Child) OR (tw:infant$ OR tw:newborn$ OR tw:neonat$ OR tw:child* OR tw:kid OR tw:toddler$ OR tw:adolescent* OR tw:teen OR tw:Youth$ OR tw:pubert$ OR tw:paediatric$ OR tw:pediatric$ OR tw:school$ OR tw:kindergar* OR tw:patient$))  2. (tw:rifampicin OR tw:rifampin OR mh:rifampin OR mh:isoniazid OR tw:isoniazid OR mh:ethambutol OR tw:ethambutol OR mh:pyrazinamide OR tw:pyrazinamide$ OR tw:antitubercul$ OR mh:"antitubercular antibiotics" OR tw:antimycobacterial$)  3. (((((tw:concentration$ OR tw:level* OR tw:monitoring) AND (tw:serum OR tw:plasma)) OR tw:pharmacokinetic$ OR tw:"therapeutic drug monitoring")))  4. ((ti:"case report" OR pt:"case reports" OR tw:editorial OR ti:letter OR ti:animal$ OR mh:"Laboratory Animals" OR mh:"Animal Experimentation" OR mh:Rodentia OR ti:rats OR ti:rat OR ti:mouse OR ti:mice))  5. ((#1 AND #2) AND #3) NOT #4 | ( ( ((mh:Infant OR mh:Adolescent OR mh:Child) OR (tw:infant$ OR tw:newborn$ OR tw:neonat$ OR tw:child* OR tw:kid OR tw:toddler$ OR tw:adolescent* OR tw:teen OR tw:Youth$ OR tw:pubert$ OR tw:paediatric$ OR tw:pediatric$ OR tw:school$ OR tw:kindergar* OR tw:patient$)) AND (tw:rifampicin OR tw:rifampin OR mh:rifampin OR mh:isoniazid OR tw:isoniazid OR mh:ethambutol OR tw:ethambutol OR mh:pyrazinamide OR tw:pyrazinamide$ OR tw:antitubercul$ OR mh:"antitubercular antibiotics" OR tw:antimycobacterial$) ) AND (((((tw:concentration$ OR tw:level* OR tw:monitoring ) AND (tw:serum OR tw:plasma)) OR tw:pharmacokinetic$ OR tw:"therapeutic drug monitoring"))) ) NOT ( ((ti:"case report" OR pt:"case reports" OR tw:editorial OR ti:letter OR ti:animal$ OR mh:"Laboratory Animals" OR mh:"Animal Experimentation" OR mh:Rodentia OR ti:rats OR ti:rat OR ti:mouse OR ti:mice))) |
